# Longitudinal sequencing and variant detection of SARS-CoV-2 across Southern California wastewater from April 2020 – August 2021

**DOI:** 10.1101/2023.04.14.23288559

**Authors:** Jason A. Rothman, Andrew Saghir, Amity G. Zimmer-Faust, Kylie Langlois, Joshua A. Steele, John F. Griffith, Katrine L. Whiteson

## Abstract

Wastewater based epidemiology (WBE) is a useful method to detect pathogen prevalence and may serve to effectively monitor diseases at a broad scale. WBE has been used throughout the COVID-19 pandemic to track localized and population-level disease burden through the quantification of SARS-CoV-2 RNA present in wastewater. Aside from case load estimation, WBE is being used to assay viral genomic diversity and the emergence of potential SARS-CoV-2 variants.

Here, we present a study in which we sequenced RNA extracted from sewage influent samples obtained from eight wastewater treatment plants representing 16 million people in Southern California over April 2020 – August 2021. We sequenced SARS-CoV-2 with two methods: Illumina Respiratory Virus Enrichment and metatranscriptomic sequencing (N = 269), and QIAseq SARS-CoV-2 tiled amplicon sequencing (N = 95). We were able to classify SARS-CoV-2 reads into lineages and sublineages that approximated several named variants across a full year, and we identified a diversity of single nucleotide variants (SNVs) of which many are putatively novel SNVs, and SNVs of unknown potential function and prevalence. Through our retrospective study, we also show that several sublineages of SARS-CoV-2 were detected in wastewater up to several months before clinical detection, which may assist in the prediction of future Variants of Concern. Lastly, we show that sublineage diversity was similar between wastewater treatment plants across Southern California, and that diversity changed by sampling month indicating that WBE is effective across megaregions.

As the COVID-19 pandemic moves to new phases, and additional SARS-CoV-2 variants emerge, the ongoing monitoring of wastewater is important to understand local and population-level dynamics of the virus. Our study shows the potential of WBE to detect SARS-CoV-2 variants throughout Southern California’s wastewater and track the diversity of viral SNVs and strains in urban and suburban locations. These results will aid in our ability to monitor the evolutionary potential of SARS-CoV-2 and help understand circulating SNVs to further combat COVID-19.

## Introduction

The COVID-19 pandemic has had a profound impact on the human population, causing over 600 million cases of disease and more than 6 million human deaths worldwide (1). Caused by the emergence of the +ssRNA “severe acute respiratory syndrome coronavirus 2” (2), COVID-19 has caused public health to react and respond in novel ways to track the spread of disease (3–5). One of these unexpected responses has been through the use of Wastewater-based epidemiology (WBE) to monitor SARS-CoV-2 viral loads in wastewater throughout the COVID-19 pandemic (4, 6, 7). As part of a worldwide effort to combat COVID-19, a massive assemblage of epidemiologists has developed methods and analyses for the examination of SARS-CoV-2 viral material in wastewater to track the spread and approximate cases of COVID-19 (4, 8–10). While direct sampling from patients is the most definitive method of COVID-19 diagnosis (11, 12), it has been shown that clinical testing has probably undercounted the true number of cases (3, 13–15). This inaccuracy in case counts is likely due to a combination of supplies issues, inability or reluctance to be tested for COVID-19, asymptomatic disease, and unreported at-home-testing (3, 13, 14). As a partner to traditional public health responses, WBE has shown to be a valuable tool in predicting and assaying case counts across populations both small and large (4, 7, 16, 17). Though WBE has shown to be a vital component of the world’s fight against COVID-19, its major method of RT-qPCR on extracted RNA from wastewater samples is only able to quantify viral loads and cannot monitor the evolution of SARS-CoV-2 and the resulting viral variants.

The SARS-CoV-2 virus has mutated many times since its original genomic description, representing ongoing evolution as the COVID-19 pandemic progresses (18–21). The WHO and PANGO Network monitor these mutations broadly classifying variants into Variants of Concern (i.e. Alpha, Beta, Gamma, Delta, Omicron), Variants of Interest (i.e. Epsilon, Zeta, Eta, Mu), or Variants under Monitoring, along with lineage designations based on phylogenetics (i.e. B.1.351, B.1.1.529) (20–23). In many cases, SARS-CoV-2 variants may possess mutations that confer phenotypic changes into the COVID-19 disease, such as increased transmissibility or antibody escape (18, 24). While these variants often contain numerous mutations, single nucleotide polymorphisms (SNPs) have been shown to occur across the SARS-CoV-2 genome, (known as Single Nucleotide Variants; SNVs) representing mutational events that often have unknown functional or evolutionary consequences (17, 25–28). Currently, direct sampling from COVID-positive patients is the gold standard for SARS-CoV-2 sequencing but remains limited by the logistics required to administer tests and only allows for the sequencing of virus from one patient at a time. Likewise, single-isolate patient sequencing likely misses rare SARS-CoV-2 variants (29), or those infecting non-human hosts, which may serve as undetected reservoirs for SNVs (30). By sequencing SARS-CoV-2 from wastewater, we can capture circulating variants/SNVs across wide areas, which provides a composite sample representing large populations and may detect SNVs before standard medical sampling (17, 25, 29, 31).

There are many challenges to sequencing SARS-CoV-2 from wastewater samples (32–34). As a matrix of industrial, agricultural, and human-borne wastes, wastewater often contains a variety of detergents and other compounds that serve as PCR inhibitors and likely degrade viral particles (29, 32, 35, 36). Similarly, SARS-CoV-2 is often at a low viral load, and the virus detected in wastewater is almost certainly fragmented, making sequencing difficult due to an inability to cover an entire genome in one assay (17, 37–39). Sequencing methods have been developed to address these challenges, such as viral enrichment, targeted amplification of viral regions, and various RNA extraction protocols, but many of these methods are designed for clinical samples, so wastewater analyses remain technically difficult to accurately conduct (16, 17, 40, 41). In order to increase our confidence in SARS-CoV-2 variant analyses, we used two sequencing library preparation methods on wastewater samples: The Illumina Respiratory Virus Oligonucleotide Panel, which enriches for respiratory virus nucleic acids before sequencing, and the QIAseq SARS-CoV-2 Primer Panel, which uses 200 PCR primer sets to amplify the entire SARS-CoV-2 genome.

Sequencing SARS-CoV-2 obtained from wastewater is a critical component to monitoring the ongoing COVID-19 pandemic (4). Here we present a study in which we used metatranscriptomic sequencing and two methods of library preparation (Illumina Respiratory Virus Oligonucleotide Panel or QIAseq SARS-CoV-2 Primer Panel) to identify SNVs, clades, and sublineages of SARS-CoV-2 on 317 influent wastewater samples. These samples were obtained from eight WTPs across Southern California from April 2020 – August 2021 and represent the collective wastewater of approximately 16 million residents. We investigated several lines of inquiry through our study: First, what RNA viruses are represented in our samples? Second, what clades and sublineages of SARS-CoV-2 were present in Southern California’s wastewater, and can we detect variants of concern in wastewater? Third, what SNVs were present in Southern California wastewater, and can we detect these variants with both library preparation methods? Lastly, does wastewater sequencing allow for early detection of variants before clinical sequencing?

## Materials and Methods

### Sample collection and handling

We previously reported the sample collection and handling procedure in Rothman et al 2021 (17) and Rothman et al 2022 (42). Briefly, we collected 317 1-liter 24-hour composite influent wastewater samples by autosampler at eight WTPs across Southern California between April 2020 – August 2021 (Table 1). We aliquoted and stored 50 mL of sample at 4 °C until processing.

**Table 1:**
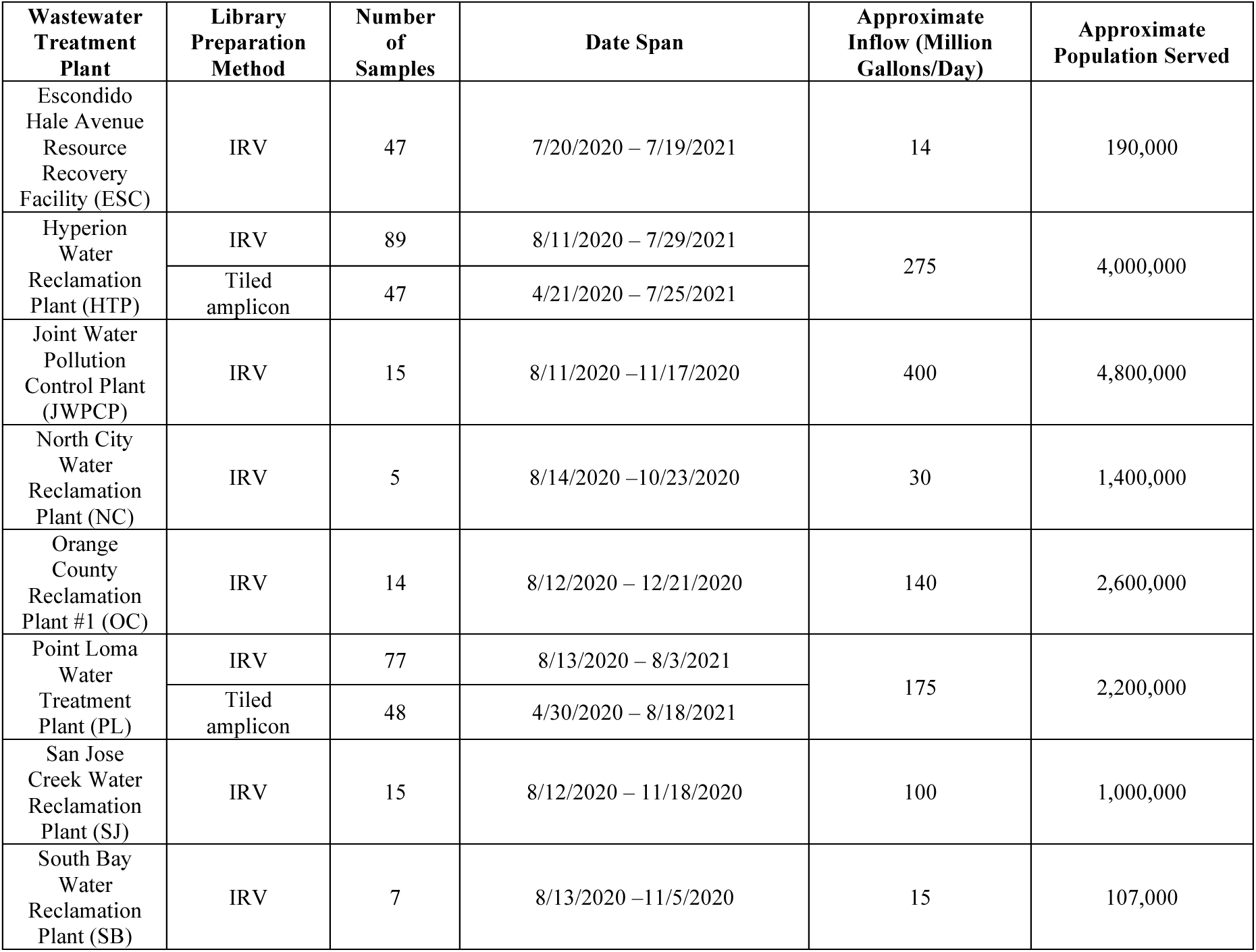
Sample quantities, date spans of collection, and approximate influent flow and served population. WTP names include the abbreviations used throughout the study, and “IRV” denotes Illumina Respiratory Virus Enrichment library preparation.

| Wastewater Treatment Plant | Library Preparation Method | Number of Samples | Date Span | Approximate Inflow (Million Gallons/Day) | Approximate Population Served |
| --- | --- | --- | --- | --- | --- |
| Escondido Hale Avenue Resource Recovery Facility (ESC) | IRV | 47 | 7/20/2020 – 7/19/2021 | 14 | 190,000 |
| Hyperion Water Reclamation Plant (HTP) | IRV | 89 | 8/11/2020 – 7/29/2021 | 275 | 4,000,000 |
|  | Tiled amplicon | 47 | 4/21/2020 – 7/25/2021 |  |  |
| Joint Water Pollution Control Plant (JWPCP) | IRV | 15 | 8/11/2020 – 11/17/2020 | 400 | 4,800,000 |
| North City Water Reclamation Plant (NC) | IRV | 5 | 8/14/2020 – 10/23/2020 | 30 | 1,400,000 |
| Orange County Reclamation Plant #1 (OC) | IRV | 14 | 8/12/2020 – 12/21/2020 | 140 | 2,600,000 |
| Point Loma Water Treatment Plant (PL) | IRV | 77 | 8/13/2020 – 8/3/2021 | 175 | 2,200,000 |
|  | Tiled amplicon | 48 | 4/30/2020 – 8/18/2021 |  |  |
| San Jose Creek Water Reclamation Plant (SJ) | IRV | 15 | 8/12/2020 – 11/18/2020 | 100 | 1,000,000 |
| South Bay Water Reclamation Plant (SB) | IRV | 7 | 8/13/2020 – 11/5/2020 | 15 | 107,000 |

### Wastewater sample RNA extraction

We used two separate RNA extraction and library preparation protocols for the samples in this study. For one set (N = 269), we used a protocol based on Crits-Christoph 2021 (25) and Rothman 2021 (17), and as the samples’ RNA was extracted for both this study and Rothman et al 2022 (42), we refer to that paper for detailed RNA extraction methods. Briefly, samples were pasteurized at 65 °C for 90 minutes in a water bath, filtered through 0.22-µM sterile filters (VWR, Radnor, PA), then centrifugated at 3000 xg with 10-kDa filters (MilliporeSigma, Burlington, MA) and RNA was extracted with an Invitrogen PureLink RNA Mini Kit plus DNase (Invitrogen, Waltham, MA). We will refer to these samples as “Illumina Respiratory Virus enriched” (IRV) henceforth.

A second set of RNA extractions was carried out (N = 95) with a different extraction and library preparation protocol based on Rothman 2021 (17) and Steele 2021 (34). Briefly, we added 25 mM MgCl_2_ to 20 mL of wastewater, then acidified the samples to pH < 3.5 with HCl. We then transferred the mixture to a cellulose ester membrane (type HA; Millipore, Bedford, MA) then bead bashed the filters in preloaded 2 mL ZR BashingBead lysis tubes (Zymo, Irvine, CA) for 1 minute. Lastly, we extracted total nucleic acids with a NucliSENS extraction kit with magnetic bead capture following the supplied protocol (bioMérieux, Durham, NC). Libraries for these samples were then prepared as follows and will be referred to as “tiled amplicons.”

### Sequencing library preparation

All sample library preparation and sequencing steps were carried out by the University of California Irvine Genomics High Throughput Facility (GHTF). The GHTF prepared IRV-enriched libraries with the Illumina Respiratory Virus Oligonucleotide panel paired with an Illumina RNA prep with enrichment kit (Illumina, San Diego, CA) following the manufacturer’s protocol. The tiled amplicon libraries were prepared by using the QIAseq SARS-CoV-2 Primer Panel paired with a QIAseq FX DNA Library UDI kit (Qiagen, Germantown, MD) and using the manufacturer’s protocol. The GHTF sequenced the resulting paired-end libraries as either 2 x 100 bp or 2 x 150 bp (supplemental file SF1) on an Illumina NovaSeq 6000 with an S4 300 cycling kit and sent the data as demultiplexed FASTQ files.

### Bioinformatics and sequence data processing

All data processing was conducted on the UCI High Performance Community Computing Cluster (HPC3). We removed sequencing adapter sequences and low-quality bases with the BBTools software “bbduk” (43). We subsequently marked sequencing duplicates with Picard toolkit “MarkDuplicates” (44), removed reads mapping to the HG38 human genome with Bowtie2 (45), then used Kraken2 (46) and Bracken (47) to taxonomically classify our reads for reporting purposes and plotted those relative abundances as a stacked bar plot with the R package “ggplot2” (48, 49).

Once we had removed the human reads, we aligned the reads to the SARS-CoV-2 Hu-1 reference strain (50) with Bowtie2, then sorted and indexed the resulting bam files with “samtools” (51). We used iVar (52) with default settings to trim off QIAseq primer sequences and call single nucleotide variants (SNVs) with a Fisher’s exact test of P < 0.05 for as compared to the reference strain (supplemental file SF1). Subsequently, we used Freyja (29) to assign SARS-CoV-2 lineage and sublineage identities to the alignments using the UShER phylogeny (53) and then de-mix the clades and sublineages within each sample to calculate approximate relative abundances. As we wanted to compare the results from IRV-enriched libraries to tiled amplicon libraries, we also used the iVar/Freyja pipeline to call SNVs and assign lineages/sublineages to these libraries even though there were no true primers to remove.

To compare our wastewater sequence data to clinical sequencing, we obtained the date of sublineage detection and reported genomes from PANGO, GISAID, and the California Health and Human Services Agency (20, 21, 54, 55) and associated these dates to our wastewater sampling dates. Due to the longitudinal nature of our data, we compared variant lineage abundances (as counts per million) over time with MaAsLin2 (56) where we had yearlong data, using WTP and sequencing batch as random effects. Likewise, we reported SNVs from 68 samples with detectible SARS-CoV-2 in Rothman 2021 (17), but here we reanalyzed the data and present the new results for consistency with our new methods.

We investigated SARS-CoV-2 sublineage alpha diversity through Kruskal-Wallis testing and beta diversity through Adonis PERMANOVA testing with the R package “vegan” (57) and measured the change in diversity with linear mixed effects regression models (LMERs) with the R package “lmerTest” (58) using both WTP and sequencing batch as random effects. Lastly, we plotted all of the data with the R packages “ggplot2,” “ggrepel” (59), “Rcartocolor” (60), and “Patchwork” (61).

### Data availability

Representative analyses scripts and code are available at https://github.com/jasonarothman/wastewater_sarscov2_apr20_aug21, and raw sequencing files have been deposited at the NCBI Sequence Read Archive under accession number PRJNA729801. SARS-CoV-2 lineage assignments and SNV calls are available in supplemental file SF1, the California Health and Human Service Agency COVID-19 Variants Dataset (55), and GISAID data are available by request from GISAID (https://gisaid.org/) (54) per their terms of use.

## Results

We used two library preparation techniques on our samples (IRV-enriched and tiled amplicon), so we report the summary statistics separately below. For IRV-enriched samples, we sequenced 548,883,572 nonhuman quality-filtered paired-end reads (average = 1,020,230, range = 9,910 – 8,243,363) across 269 samples. We taxonomically classified an average of 58.8% of reads (range = 8.9 – 85.4%) of which an average of 9.4% of overall reads were viral (range = 0.1 – 53.7%). Of total viruses, 2,281,212 reads (6.0%) mapped to SARS-CoV-2. Regarding tiled-amplicon samples, we sequenced 1,074,798,497 nonhuman quality-filtered paired-end reads (average = 11,313,668, range = 8,619,210 – 14,384,197) across 95 samples. We classified an average of 47.2% of reads (range = 35.9 – 66.7%) of which an average of 4.5% were viral (range = < 0.01% - 41.6%). Of total tiled-amplicon prepared viruses, 47,427,550 reads (99.6%) mapped to SARS-CoV-2 (Fig. 1).

**Figure 1:**
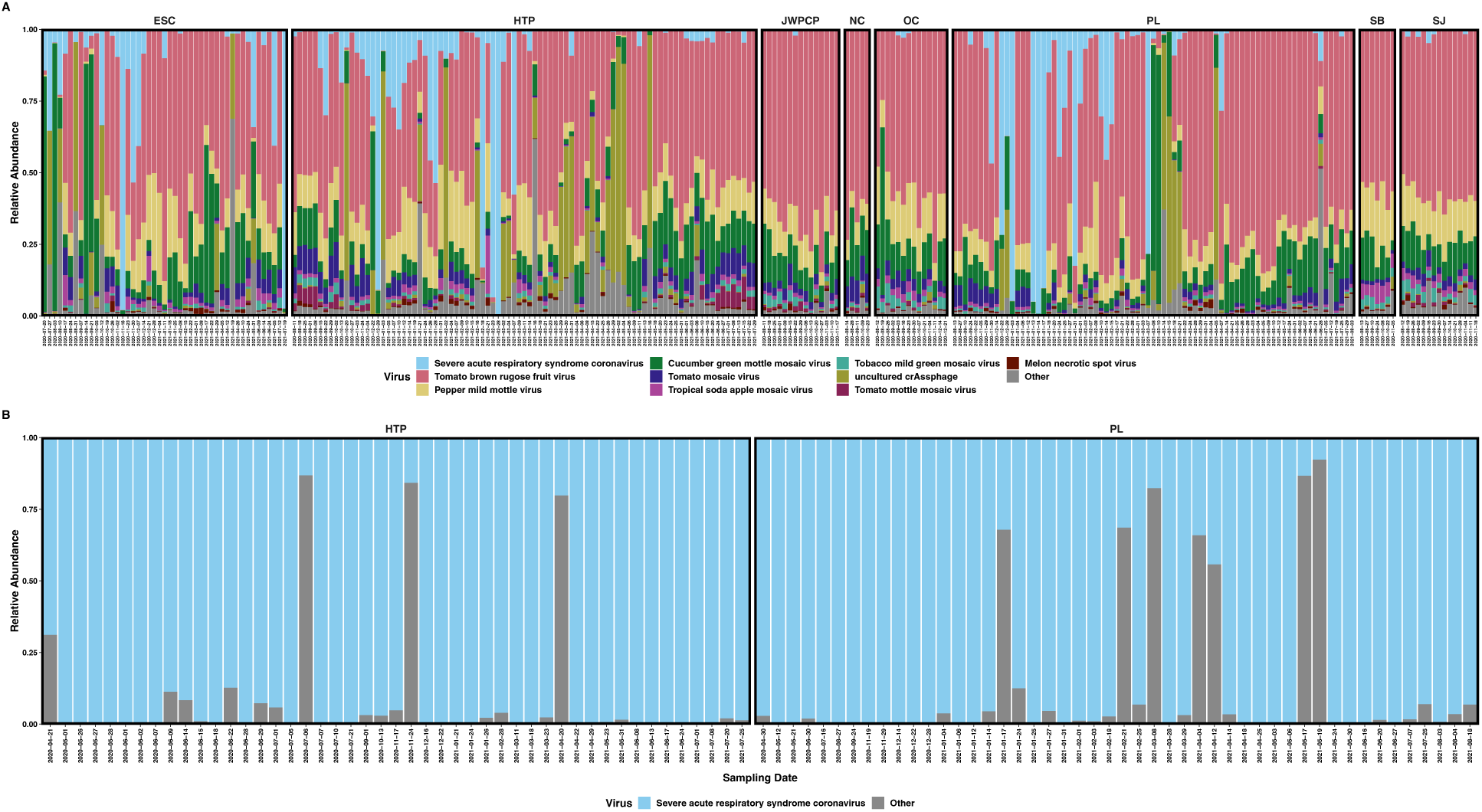
Stacked bar plots showing the relative abundances of RNA reads mapping to A) the top 10 most proportionally abundant viruses plus all others in respiratory virus-enriched libraries and B) SARS-CoV-2 plus other viruses in tiled-amplicon libraries. Plots are faceted by WTP and labeled with sampling date.

We obtained broad SARS-CoV-2 genome coverage with both sequencing approaches. When considering all samples together, IRV-prepared libraries covered 99.92% of the SARS-CoV-2 genome at a mean sequencing depth of only 2x at each base position. Tiled amplicon library preparation had both wider coverage and higher sequencing depth, with these libraries covering 99.95% of the genome at a mean depth of 76 reads per base (Fig. S1).

Because SARS-CoV-2 was well-represented in our samples, we could classify reads mapping to SARS-CoV-2 through UShER SARS-CoV-2 barcoding and de-mixing to approximate relative abundances with Freyja. We were able to classify many of the mapped reads to specific named Variants of Interest (VOIs) and Variants of Concern (VOCs) (both currently circulating and historically significant) along with other sublineages of SARS-CoV-2 that do not correspond to a VOI/VOC. Within tiled-amplicon samples that had classifiable reads (N = 90), these “top 10” most proportionally abundant clades were Alpha (x̄ = 3.3%, range = 0 – 98.8%), Beta (x̄ = 0.23%, range = 0 – 17.2%), Delta (x̄ = 16.0%, range 0 – 100%), Epsilon (x̄ = 4.6%, range = 0 – 98.9%), Gamma (x̄ = 0.02%, range = 0 – 14.3%), Iota (x̄ = 0.02%, range = 0 – 0.7%), Lambda (x̄ = 1.1%, range = 0 – 96.8%), Mu (x̄ = 0.1%, range = 0 – 8.6%), Zeta (x̄ = 0.01%, range = 0 – 0.3%), and all other sublineages combined (x̄ = 67.7%, range = 0 – 99.9%) (Fig. 2 and Fig. S2). While individual samples contained varying proportions of WHO clades, only the relative abundance of the Delta variant was shown to increase throughout the course of the experiment (β =2.57, P_adj_ < 0.001)

**Figure 2:**
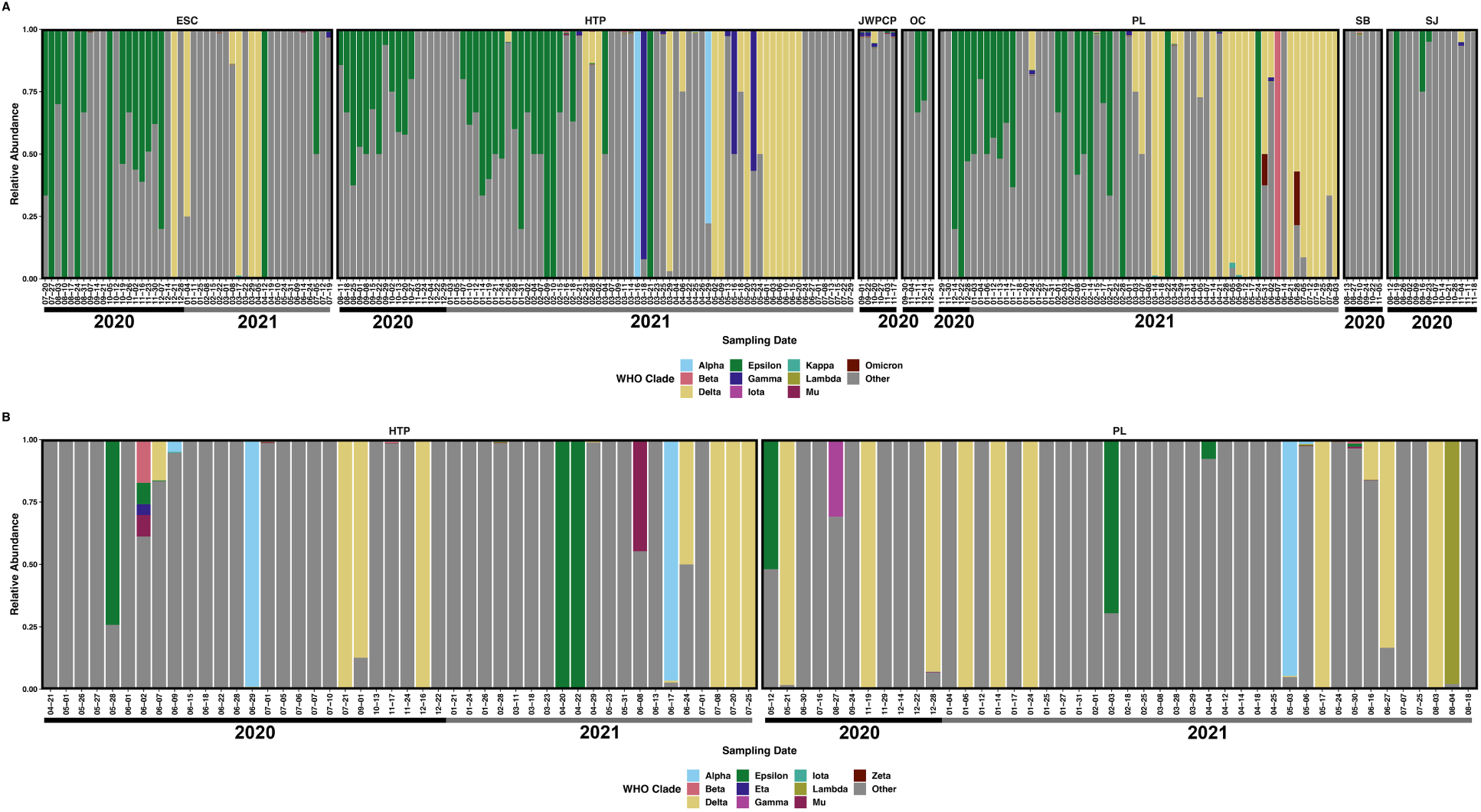
The relative proportional abundance of the ten most abundant SARS-CoV-2 lineages plus others in A) respiratory virus-enriched libraries, B) tiled-amplicon libraries faceted by WTP and labeled with sampling date. Note that one sample date from the North City Water Reclamation Plant is not shown.

Within IRV-enriched samples that had classifiable reads (N = 219), the “top 10” most proportionally abundant SARS-CoV-2 clades were largely similar to tiled-amplicon samples. These clades were Alpha (x̄ = 0.86%, range = 0 – 100%), Beta (x̄ = 0.50%, range = 0 – 100%), Delta (x̄ = 15.8%, range = 0 – 100%), Epsilon (x̄ = 17.8%, range = 0 – 100%), Gamma (x̄ = 1.0%, range = 0 – 92.2%), Iota (x̄ = 0.01%, range = 0 – 0.30%), Kappa (x̄ = 0.03%, range = 0 – 2.1%), Lambda (x̄ = 0.02%, range = 0 – 0.4%), Mu (x̄ = 0.02%, range = 0 – 0.7%), Omicron (x̄ = 0.15%, range = 0 – 18.9%), and all other sublineages combined (x̄ = 62.2%, range = 0 – 100%) (Fig. 2 and Fig. S3). Similar to tiled-amplicon results, only the relative abundance of Delta was shown to increase throughout the course of the experiment (only samples from ESC, HTP, and PL WTPs; β = 1.54, P_adj_ = 0.002). As shown above, most of the SARS-CoV-2 reads obtained from either library preparation method were not part of a named VOI/VOC or were merely identified as SARS-CoV-2 without a confident lineage classification.

In addition to large, overarching SARS-CoV-2 clades (i.e. VOI/VOCs), we often classified reads to a named PANGO sublineage, and we detected 1,221 unique sublineages at greater than 0.1% proportional abundance, with substantial detection overlap between tiled-amplicon and IRV sequencing approaches (1,215 and 1,221 named sublineages respectively, supplemental file SF1). We often detected the presence of SARS-CoV-2 sublineages in wastewater before clinical sequencing reported detection: Tiled-amplicon sequencing detected 515 (42.7%) in samples before clinical sequencing, in some cases by as much as several months, and IRV sequencing detected 364 (30%) before clinical reports, again often with substantial lead-time as above (Fig. 3).

**Figure 3:**
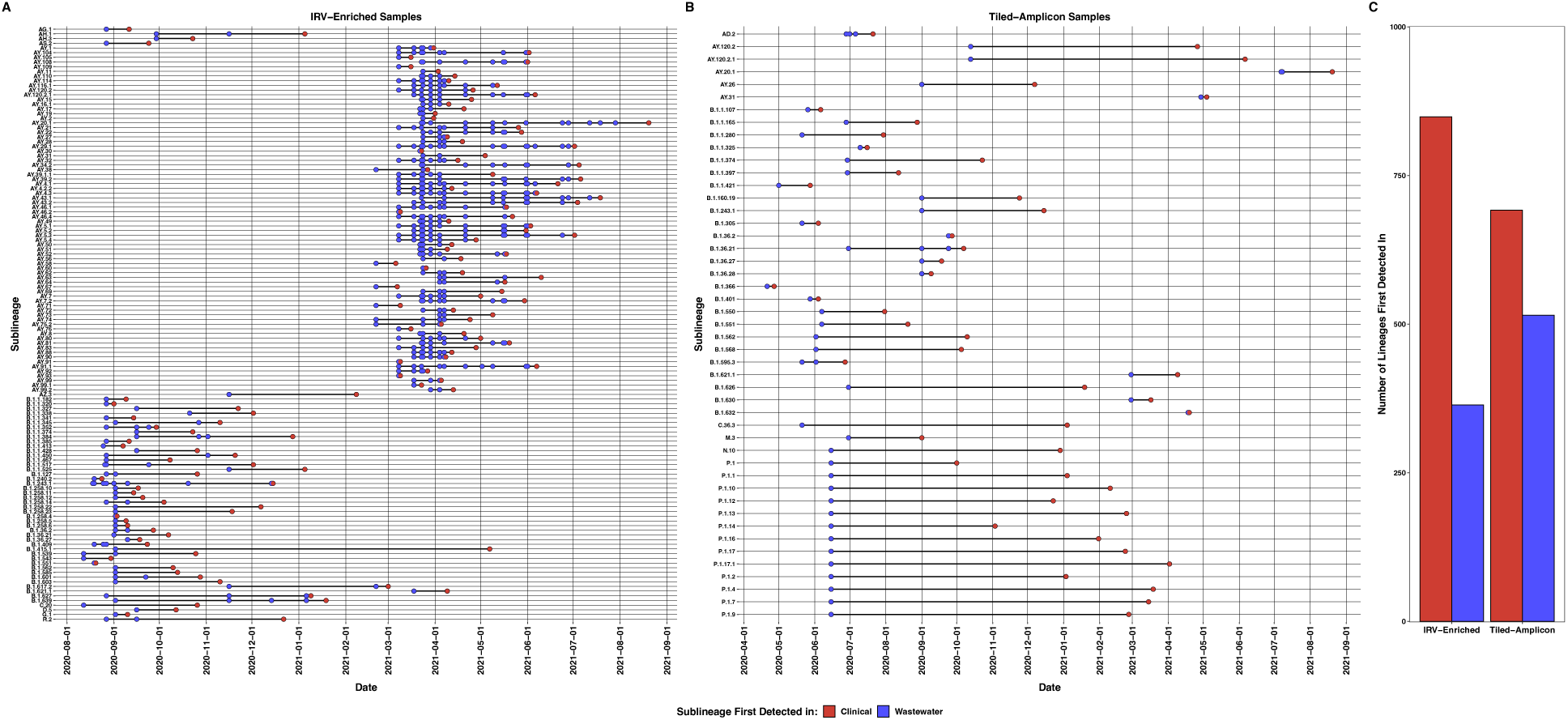
SARS-CoV-2 sublineages at greater than 0.2% relative abundance (for plot visibility) first detected in wastewater samples in A) respiratory virus-enriched libraries (IRV) and B) tiled-amplicon libraries by date. Panel C denotes the total number of SARS-CoV-2 sublineages first detected by our wastewater sequencing or clinical samples by IRV or tiled-amplicon libraries respectively without the relative abundance cutoff.

We examined the diversity of SARS-CoV-2 subclades at greater than 0.01% relative abundance where we had long term samples for both IRV-enriched (Escondido, Hyperion, and Point Loma WTPs, N = 187) and tiled amplicon (Hyperion and Point Loma WTPs, N = 90). The sublineage alpha diversity of IRV-enriched samples did not differ between WTPs (H_(2)_ = 2.1, P = 0.34) or month (H_(13)_ = 18.8, P = 0.13), nor did it differ over numerical time (t = 0.22, P = 0.82). Beta diversity of the sublineages was not different between WTPs (R^2^ = 0.01, P = 0.07), but differed between months (R^2^ = 0.15, P < 0.001) and sequencing batches (R^2^ = 0.03, P < 0.001) with no interaction between month and WTP (R^2^ = 0.10, P < 0.12), and did not change over numerical time (t = 0.15, P = 0.88) (Fig. 4).

**Figure 4:**
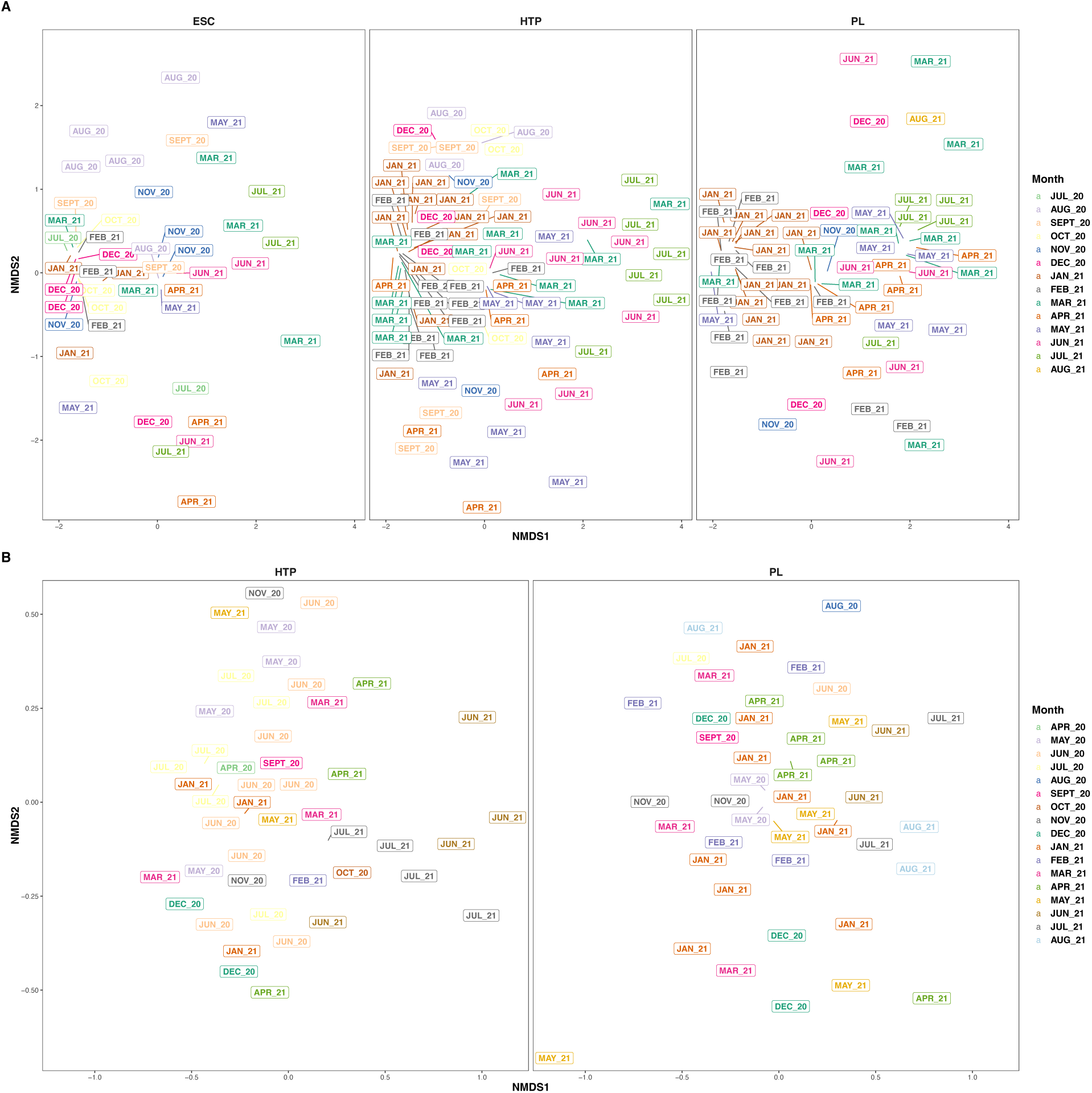
Non-metric multidimensional scaling (NMDS) ordinations of the Bray-Curtis dissimilarities of SARS-CoV-2 sublineages faceted by water treatment plant for A) respiratory virus-enriched (IRV) and B) tiled-amplicon libraries. SARS-CoV-2 sublineages did not significantly differ between WTPs (PERMANOVA [IRV: P = 0.07, R^2^ = 0.01], [tiled-amplicon: P = 0.58, R^2^ = 0.01]) but differed by calendar month (PERMANOVA [IRV: P < 0.001, R^2^ = 0.15], [tiled-amplicon: P < 0.001, R^2^ = 0.21]). Color and plot labels denote sampling month, and only WTPs with yearlong data are included.

We analyzed the tiled-amplicon samples in the same fashion as above, and did not find a difference in sublineage alpha diversity between WTP (H_(1)_ = 0.04, P = 0.84) or calendar month (H_(16)_ = 11.2, P = 0.80), and diversity remained constant over numerical time (t = 0.41, P = 0.68). We observed a difference in beta diversity by month (R^2^ = 0.21, P < 0.001), but not between WTPs (R^2^ = 0.01, P = 0.58), sequencing batches (R^2^ = 0.01, P = 0.20) or an interaction between WTP and month (R^2^ = 0.13, P = 0.19), nor by numerical time (t = −1.8, P = 0.07) (Fig. 4).

As we were unable to collect samples from all eight WTPs for the full year, we also analyzed subclade diversity during months with the broadest WTP coverage (August – November 2020 without WTP “NC” as there was only one sample, N = 61). There was no difference in sublineage alpha diversity between WTPs (H_(6)_ = 11.9, P = 0.07), months (H_(3)_ = 2.5, P = 0.47), or numerical time (t = −1.4, P = 0.17). There was a significant difference in sublineage beta diversity between WTPs (R^2^ = 0.15, P < 0.001), and an interaction between WTP and month (R^2^ = 0.21, P < 0.027), with no differences by calendar month (R^2^ = 0.06, P = 0.09) or sequencing batch (R^2^ = 0.01, P = 0.62), or numerical time (t = −0.8, P = 0.41).

At a more granular level, were also tabulated SARS-CoV-2 Single Nucleotide Variants (SNVs) throughout our samples (supplemental file SF1). Combined, IRV-enriched and tiled-amplicon sample preparation methods detected 2,871 SARS-CoV-2 SNVs across the genome, with each approach capturing a different number of SNVs, and in many cases, different genomic locations (Fig. 5). IRV-enriched samples contained 1,212 SNVs, most being found only once (1,071) or twice (83), however we often detected the same SNV multiple times at several genomic positions in separate samples (Fig. 5). For example, SNVs at nucleotide positions 23403, 241, 14408, 17014, 3037, 28272, 8947, 12878, 2597, 21600, 25563, and 28887 were each detected over 10 times across the samples. Tiled-amplicon sequencing also detected SNVs well, identifying 1,808 SNVs across the SARS-CoV-2 genome. Similar to IRV-enriched results, most SNVs were found once (1,030) or twice (254), although several SNVs were identified in multiple samples (Fig.5): SNVs located at nucleotide positions 22796, 28971, 22656, 28982, and 9864 were detected over 40 times across tiled-amplicon samples.

**Figure 5:**
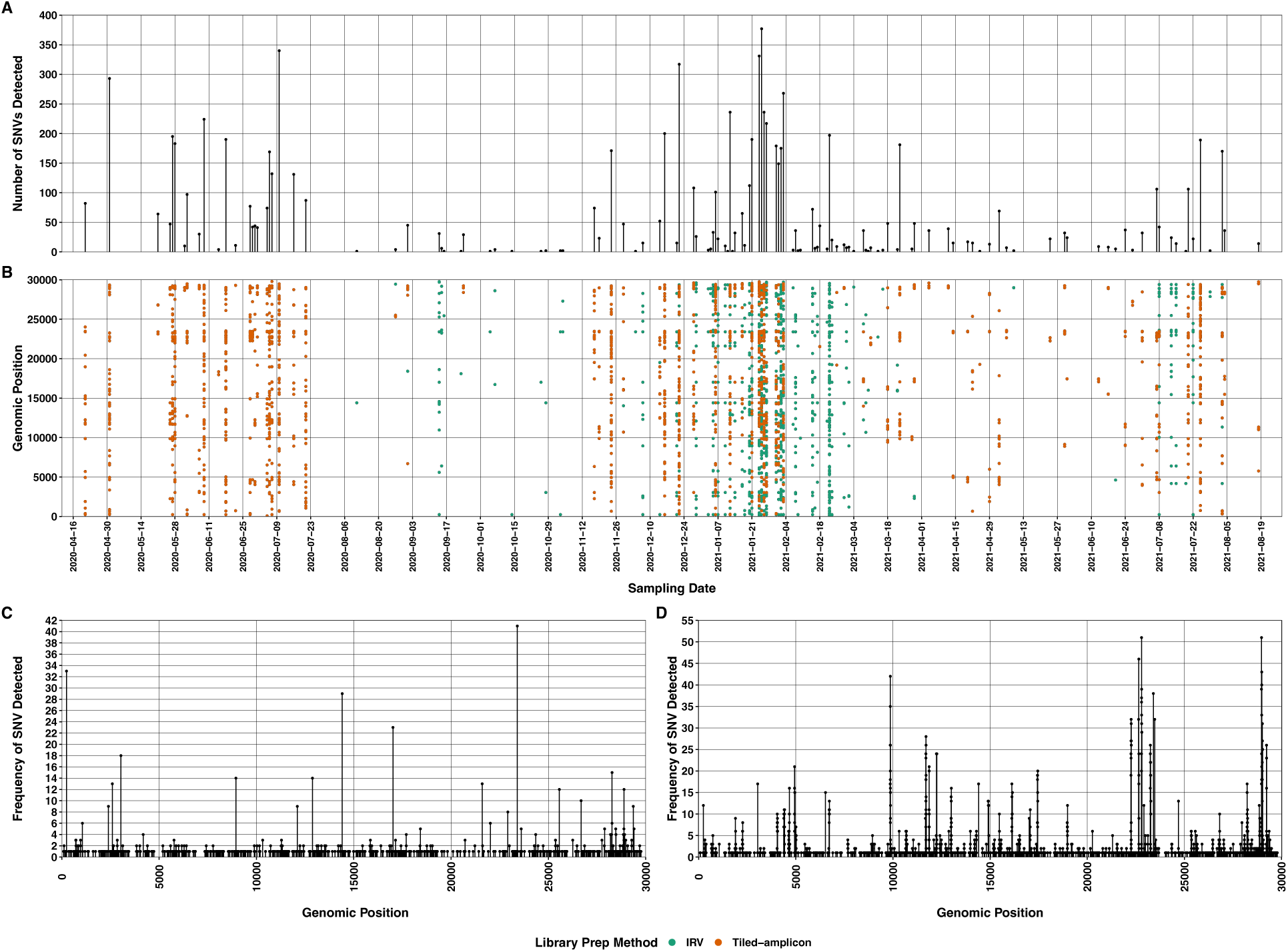
A) Number of single nucleotide variants (SNVs) detected at each sample date and B) nucleotide position across the SARS-CoV-2 genome for all samples colored by library preparation method (IRV signifies Illumina Respiratory Virus enrichment panel). Panels C and D indicate the frequency of SNVs detected at each position of the SARS-CoV-2 genome across all respiratory virus-enriched and tiled-amplicon libraries respectively.

## Discussion

Our study represents a large-scale effort to employ wastewater-based epidemiology (WBE) across a catchment area of 16 million people and supports the monitoring of SARS-CoV-2 evolution throughout the ongoing COVID-19 pandemic. Through respiratory virus-enriched and tiled-amplicon RNA sequencing approaches, we classified SARS-CoV-2 lineages and single nucleotide variants (SNVs) and could approximate VOCs/VOIs across a yearlong study of Southern California wastewater. Like other studies, we captured SARS-CoV-2 mutations across the genome and show the potential to detect sublineages and SNVs months before clinical analyses of patient samples (17, 25, 26, 29, 38). While WBE is a powerful tool - and is not subject to many of clinical sequencing’s drawbacks - we cannot use these methods to determine the exact source of SARS-CoV-2 variants (17, 25, 29, 37) and instead propose the use of WBE to monitor populations instead of individuals. Our results suggest that multi-scale sampling of individual patients, local wastewater catchments (i.e. university campuses), and WTPs can give public health agencies vital information to identify novel SARS-CoV-2 variants and predict disease spread to further combat COVID-19 (4, 10, 29).

In most samples, we could classify SARS-CoV-2 RNA fragments at multiple levels of resolution – both at the named variants (i.e. Alpha, Beta, etc) and sublineage levels (i.e. B.1.429, B.1.617.2, P.1, etc) and calculate semi-quantitative relative abundances. When considering the full year of data, sublineage diversity of SARS-CoV-2 was not different between WTPs, rather it changed monthly probably due to the similarity of proportional disease burden and proximity of San Diego and Los Angeles counties (1). As expected, our sublineage quantification was not exactly concordant with clinical sequencing data, probably due to the aggregate nature of wastewater and our composite sampling, along with the lack of clinical specimens early in the pandemic (4, 20, 21, 29, 37). We do note however that ours and clinical data agree well during the emergence of the Delta variant, suggesting that wastewater can detect the potential evolutionary replacement of lineages accurately as has been recently shown with the domination of the Omicron variant (29, 62). Similarly, we detected many SARS-CoV-2 sublineages earlier in wastewater than clinical sequencing – in some cases by several months – further supporting work indicating that WBE is useful for predicting disease load and the spread of novel variants (29, 62). Naturally, we recognize this is a retrospective study, and rely on clinical sequencing to name and prioritize the variants we sequenced in wastewater, so we suggest that public health and wastewater sequencing be used in tandem to carefully monitor the evolutionary potential of SARS-CoV-2 (4).

Similar to previous work, we detected thousands of single nucleotide variants (SNVs) across samples and sequenced putatively novel or rare SNVs that have unknown function or species host (17, 25, 26, 30, 62, 63). For example, in many samples (from within or between WTPs), we detected SNVs at positions 9864, 22796, and 28971 which are exceedingly rare in public sequencing data, along with SNVs at 241, 14408, and 23403 which were common in 2020 (64, 65). Our ability to detect both low-prevalence and near-ubiquitous SNVs indicates that WBE is broadly useful for accurate SNV detection and may provide a reasonable estimate of what SNVs are circulating across populations (25, 29, 30, 62, 63). Likewise, when comparing our results to other wastewater studies, we detected variants or sublineages also reported in Nice, New York, Montana, Arizona, Northern California, Berlin, and across Austria, often at similar sampling dates, which shows that sequencing wastewater is reproducible and accurate at very large scales (25, 30, 38, 66, 67). Being that sequencing wastewater is technically challenging, we qualitatively compared two major methods of SARS-CoV-2 analysis and note that targeted amplification provided better sequencing depth and resolution, indicating its utility when presented with degraded low-titer RNA and the detergent/PCR inhibitor content of wastewater along with our harsh extraction methods (4, 17, 40). We suggest the use of targeted amplification approaches for wastewater samples, which supports previous work and method development (4, 17, 25, 29, 40, 62, 68, 69).

## Conclusions

Wastewater-based epidemiology has exploded into a worldwide endeavor and is a critical part of humanity’s response to the COVID-19 pandemic (4, 6, 8, 10). Our study demonstrates WBE’s effectiveness in monitoring SARS-CoV-2 mutations across megaregions (17, 25, 29, 30, 38), which continues to be important as novel VOCs emerge and the popularity of at-home testing reduces public health’s ability to accurately quantify COVID-19 cases (3, 13, 14). COVID-19 has demonstrated the need for scientists, wastewater agencies, and public health to work together to track the evolution and spread of SARS-CoV-2, especially in underserved areas, low population coverage, and places where the medical field is overburdened (4, 15). WBE has the potential to discover emergent diseases and should be implemented across population centers as a sentinel for the next pandemic.

## Data Availability

Raw sequencing files have been deposited at the NCBI Sequence Read Archive under accession number PRJNA729801.

https://www.ncbi.nlm.nih.gov/bioproject/PRJNA729801

## Acknowledgments

We thank the Los Angeles and Orange County Sanitation Districts, the City of San Diego Public Utilities, the City of Escondido Hale Avenue Resource Recovery Facility, and the City of Los Angeles Department of Sanitation and Environment for collecting wastewater samples. We also thank the developers of Freyja and Adélaïde Roguet for software assistance, and Seung-Ah Chung for library preparation assistance.

This research was supported by the University of California Office of the President Research Grants Program Office (award numbers R01RG3732 and R00RG2814) awarded to JAR and KLW, and a Hewitt Foundation for Biomedical Research postdoctoral fellowship to JAR. This work was made possible, in part, through access to the Genomics High Throughput Facility Shared Resource of the Cancer Center Support Grant (P30CA-062203) at the University of California, Irvine, NIH shared instrumentation grants 1S10RR025496-01, 1S10OD010794-01, and 1S10OD021718-01, and access to computing resources from the UCI High Performance Community Computing Cluster.

## Notes

### Competing Interest Statement

The authors have declared no competing interest.

