## Supplemental information for "Longitudinal sequencing and variant detection of SARS-CoV-2 across Southern California wastewater from April 2020 – August 2021"

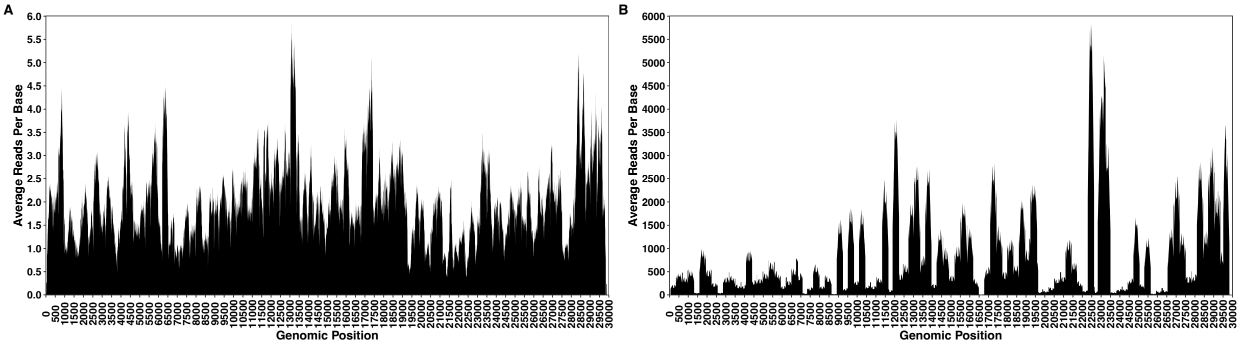


Figure S1: Area plot of the average reads per base mapped to the SARS-CoV-2 genome across all samples. Panel A represents libraries prepared with the Illumina Respiratory Virus Enrichment Panel (IRV) and Panel B represents libraries prepared for tiled amplicon sequencing.

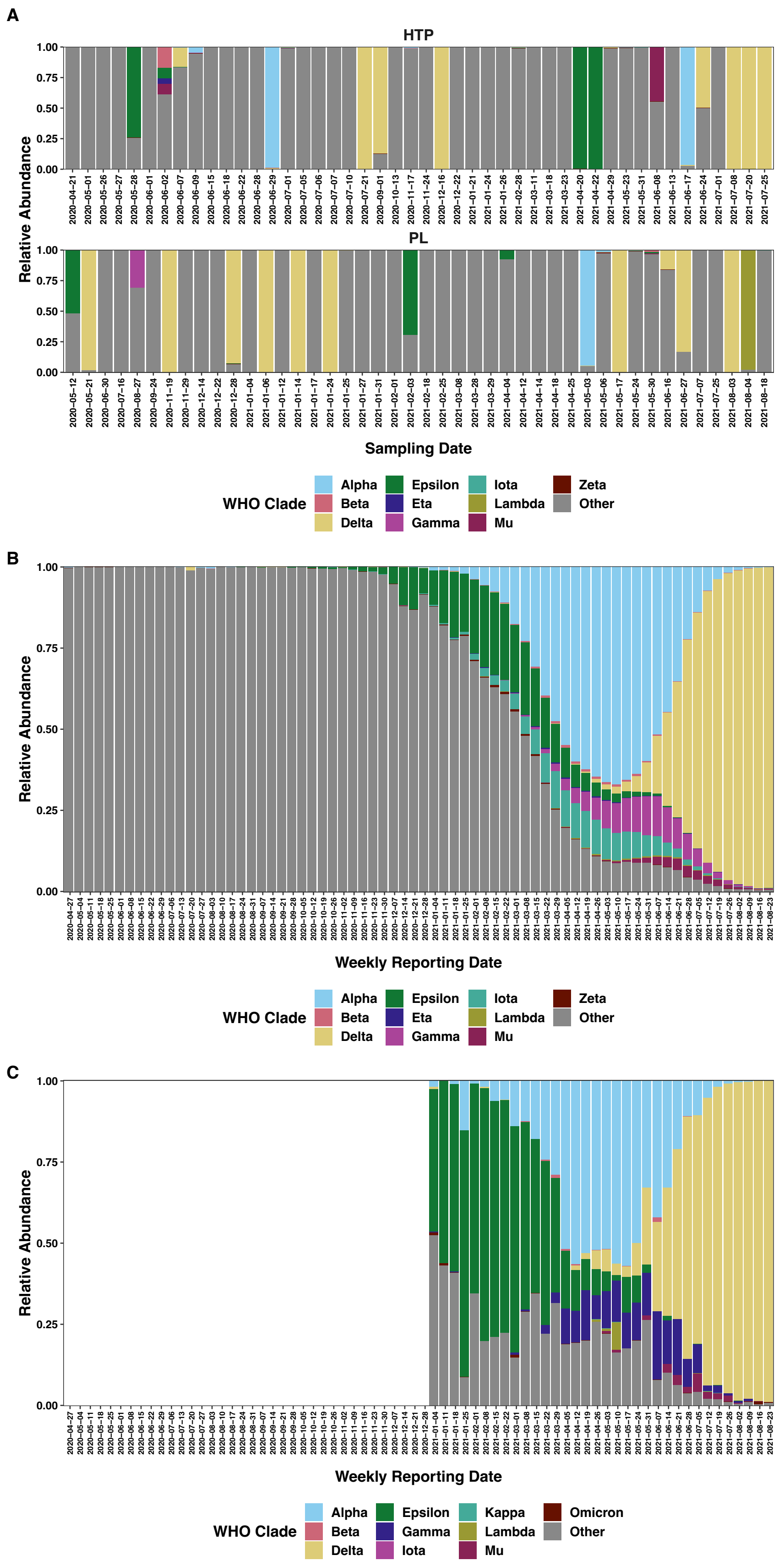
A
HTP
PL
Relative Abundance
1.00
0.75
0.50
0.25
0.00
Sampling Date
WHO Clade
Alpha
Beta
Delta
Epsilon
Eta
Gamma
Iota
Lambda
Mu
Zeta
Other
B
Relative Abundance
Weekly Reporting Date
WHO Clade
Alpha
Beta
Delta
Epsilon
Eta
Gamma
Iota
Lambda
Mu
Zeta
Other
C
Relative Abundance
Weekly Reporting Date
WHO Clade
Alpha
Beta
Delta
Epsilon
Gamma
Iota
Kappa
Lambda
Mu
Omicron
Other

Figure S2: The relative proportional abundance of the ten most abundant SARS-CoV-2 lineages plus others in A) tiled-amplicon libraries, B) weekly GISAID-reported data for clinical samples from the United States, and C) CHSS-reported clinical data. Panel A is faceted by WTP and labeled with sampling date and panel B is labeled by the aggregate of GISAID-reported data. Note that one sample date from the North City Water Reclamation Plant is not shown, and variant data from CHSS are not available before 1/1/21 so those dates are left blank.

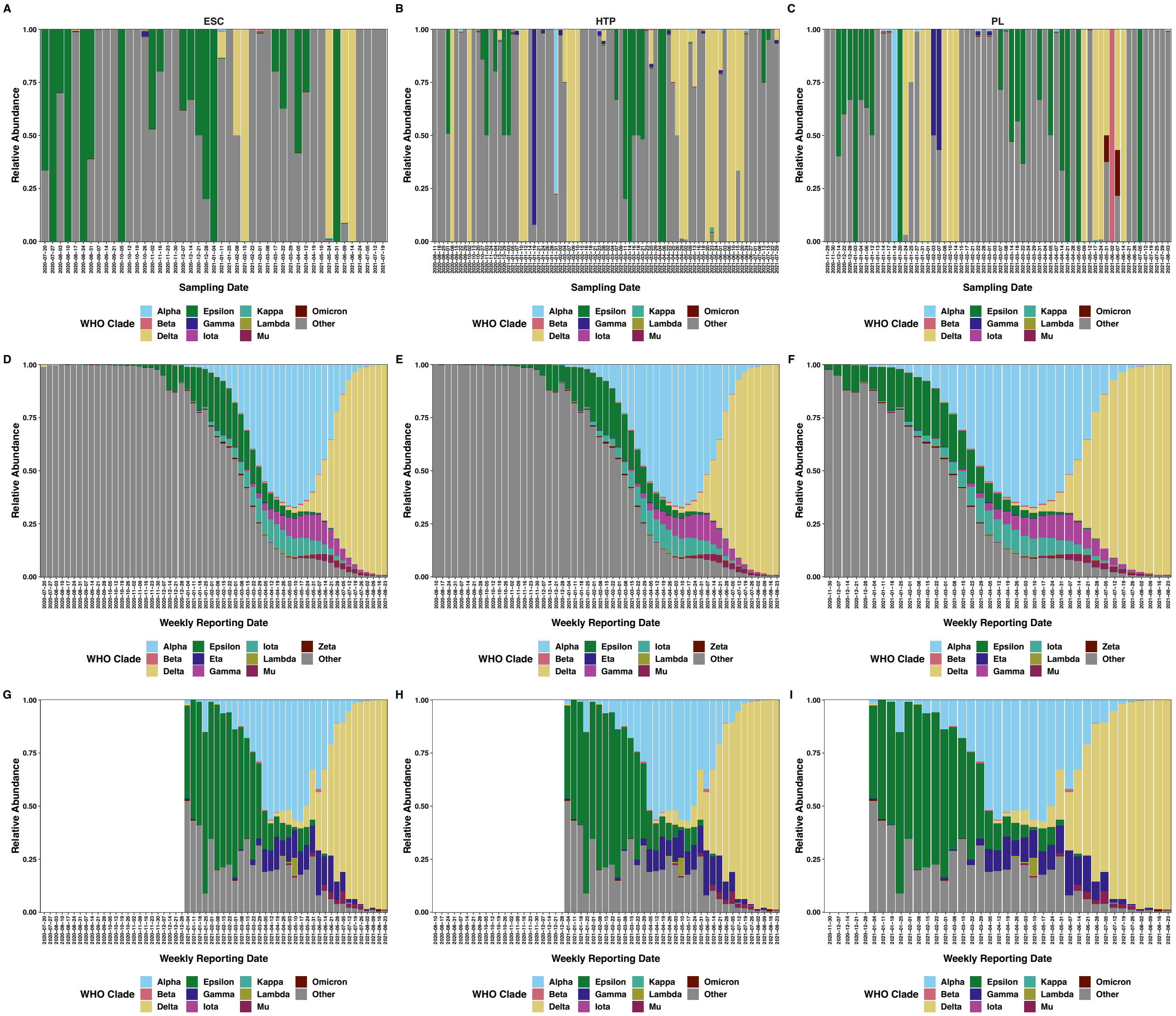


Figure S3: The relative proportional abundance of the ten most abundant SARS-CoV-2 lineages plus others in Illumina Respiratory Virus Panel enriched libraries for A) Escondido, B) Hyperion, and C) Point Loma water treatment facilities. Panels D, E, and F are weekly GISAID-reported data for clinical samples from the United States corresponding to the approximate time periods of panels A, B, and C, respectively. Panels G, H, and I are CHSS-reported clinical data corresponding to the approximate time periods of panels A, B, and C, respectively. Variant data from CHSS are not available before 1/1/21 so those dates are left blank.

Supplemental File SF1: This file contains several worksheets of information: A README sheet with detailed information, sample metadata, single nucleotide variants for IRV and tiled amplicon samples, and PANGO sublineages as output by Freyja for IRV and tiled amplicon samples.
